# Occupational Exposures to Blood and other Body Fluids among Healthcare Workers in Cameroon: A Systematic Review and Meta-analysis

**DOI:** 10.1101/2024.12.05.24318564

**Authors:** Fabrice Zobel Lekeumo Cheuyem, Christian Mouangue, Brian Ngongheh Ajong, Michel Franck Edzamba, Dilane Christian Maidey Hamadama, Chabeja Achangwa, Adama Mohamadou, Pamela Sonfack, Adidja Amani

## Abstract

**Background:** Occupational exposure to blood and body fluids has become a serious public health problem for healthcare workers and is a major risk for the transmission of bloodborne infections such as human immune-deficiency, hepatitis B, and hepatitis C viruses. It has been identified as one of the most serious issues affecting the health and well-being of health workers in most health systems especially in developing countries. The present systematic review and meta-analysis was conducted to estimate the pooled prevalence of blood and other body fluids exposures among healthcare workers in Cameroon.

**Methods:** Online platform including PubMed, Google Scholar, Cochrane Library, and Science Direct were systematically searched to collect relevant research reports. Unpublished studies in a national library were also consulted. The *I^2^* tests were used to assess the heterogeneity of the included studies. A fixed and random-effects meta-analysis model was used to estimate the lifetime and 12-month prevalence of occupational exposure to blood and other body fluids among healthcare workers in Cameroon.

**Results:** Of the 539 records identified through the database search, 15 study reports were included in the final analysis. The random-effects model showed that the estimated overall pooled prevalence of 12-month and lifetime exposure to blood and other body fluids among healthcare workers in Cameroon was 55.44% (95% Confidence Interval (CI): 41.20-69.68); (*I^2^*=97.5%; *p*<0.001) and 57.27% (95% CI: 42.43-72.10); (*I^2^*=97.7%; *p*<0.001) respectively. The highest 12-month pooled prevalence was observed in intermediate level health facilities (84.73%; 95% CI: 85.55-88.50), in Regions other than the Centre (70.87%; 95% CI: 37.26-95.13) and for studies conducted from 2017 to 2023 (65.63%; 95% CI: 45.73-83.06). The lifetime prevalence of blood and other body fluids exposures was the highest for Regions namely the North-west and South-west Regions (77.96%; 95% CI: 57.39-93.19).

**Conclusion:** Healthcare workers in Cameroon face a significant risk of occupational exposure to blood and body fluids (BBFs), with a high prevalence of exposure over their lifetime and in the past 12 months. This highlights the urgent need to enhance and implement effective occupational safety and health policies to protect healthcare workers in Cameroon.

## Background

Occupational exposure to blood and other body fluids (BBFs) poses a significant risk of transmission of blood-borne infections to healthcare workers (HCWs). Such exposures increase the risk of infection with human immunodeficiency virus (HIV), hepatitis B, and hepatitis C. In many cases, exposures occur through, splashes of blood or other body fluids into the eyes, nose, or mouth, or through non-intact skin [1,2]. It can also occur via percutaneous injuries from needlesticks and sharp objects [3]. Infectious materials include body fluids such as blood, urine, saliva, droplets, contaminated tissue, tools, equipment, surfaces and the environment [4].

Studies review indicate that occupational exposure to BBFs is one of the most serious issues affecting the health and well-being of health workers in most health systems especially in developing countries [5]. Several factors have been identified as increasing the risk of accidental exposure to BBFs. These factors include age, gender, work experience, shift work, lack of training and supervision [6,7]. The implementation of preventive measures including educational programs, correct and systematic use of personal protective equipment, and adherence to infection control and prevention guidelines, can significantly reduce the burden of occupational exposure among HCWs [8].

The reporting pattern of occupational exposure remains inadequate in many health care facilities in developing countries including Cameroon [9,10]. In addition, the lack of surveillance system leads to the underestimation of the burden of this occupational health problem [11].

Several primary reports in Cameroon revealed a high prevalence of occupational exposure to BBFs [3,4,11–13]. Currently, no report exists in Cameroon to quantify the pooled prevalence of occupational exposure to BBFs and the reporting pattern among HCWs. There is a need to document the overall burden of occupational exposure to BBFs. Such data will be useful in raising awareness and designing intervention strategies to guide the implementation of safety standards in the country’s health facilities [10,14]. The present systematic review and meta-analysis was conducted to estimate the pooled prevalence of BBFs exposure among HCWs in Cameroon.

## Methods

This systematic review was conducted in accordance with PRISMA (Preferred Reporting Items for Systematic Reviews and Meta-Analysis) guideline [15].

### Study Designs and Setting

Cameroon’s population is estimated to be approximately 28.6 million as of 2023. It covers a surface area of 472 650 km² spread across ten administrative Regions, including the Centre, Littoral, Far-North, North, Adamawa, North-West, South-West, West, East and South. The country boasts two capitals : Yaounde, located in the Centre Region, serves as the nation’s political hub, while Douala, situated in the Littoral Region, is the economic powerhouse driving the country’s growth [16]. In terms of the health system component, in 2023 Cameroon has reached the World Health Organization standard for health facility density (2.3 per 10,000 population), with 24.0 available beds per 10,000 population. The country has 11.3 qualified HCWs and reports an estimated 0.41 outpatient consultations per person per year [17]. The health system pyramid include three categories including the central, intermediate and operational level [4].

### Eligibility Criteria

This review included cross-sectional studies that reported the lifetime and/or 12-month prevalence of occupational exposure to BBFs through exposure to mucous membranes and injured skin. To be included, studies had to provide quantitative outcomes and investigate occupational exposure to BBFs as a dependent variable. Only full-text articles in English with clear objectives and methods were included. For the narrative review, additional studies investigating risk factors of occupational exposures to BBFs were also included. This review focused on studies involving HCWs in different departments of private and public healthcare settings. Specifically, the review included studies involving doctors, nurses, midwives, assistant nurses, laboratory technicians, medical students and cleaners. No restriction was applied on publication date, since there was no prior systematic review that investigated occupational exposures to BBFs in the country.

### Article Searching Strategy

A comprehensive review of various literature sources was conducted, including both published and unpublished studies, based on predetermined eligibility criteria. A systematic search of electronic databases, such as PubMed, Google Scholar, Cochrane Library, and Science Direct, was performed to identify published studies. The search strategy involved analyzing the text contained in the title and abstract of each study. A combination of keywords and MeSH (Medical Subject Headings) terms was used, employing Boolean logic operators (“AND” and “OR”) to refine the search. The keywords and MeSH terms included: “prevalence”, “occupation”, “accidental exposure”, “magnitude”, “exposure”, “occupational disease”, “accident”, “occupational exposure”, “cross infection”, “occupational hazard”, “body fluid”, “blood spill”, “blood”, “blood-borne pathogens”, “reporting”, “management”, “post-exposure prophylaxis”, “blood-borne infection”, “healthcare workers”, “health workers”, “health-care workers”, “health personnel”, “risk factor”, “circumstance”, “factor favoring”, “medical personnel” and “Cameroon”. To ensure a comprehensive search, manual searching was conducted to identify additional published articles not indexed in electronic databases. Unpublished studies were search at the University of Yaounde 1 library. Furthermore, the reference lists of identified studies were scanned to achieve literature saturation. The last search was conducted on 01 November 2024.

### Data Extraction and Quality Assessment

A standardized Microsoft Office Excel 2019 form was used to extract relevant data from the selected studies, including name of primary author, study year, region, study design, setting, study population, sample size, response rate, 12-month and lifetime prevalence of exposure to BBFs among HCWs and reporting rate. All data were extracted from the eligible articles. Three authors critically assessed each article for relevance and quality. Disagreement between reviewers were resolved by discussion. The quality of the included studies was assessed using the Joanna Briggs Institute quality assessment tool for prevalence studies [18]. Nine parameters were used to assess the risk of bias for each study, including appropriateness of sampling frame, appropriate sampling technique, adequate sample size, description of study subjects and setting, sufficient data analysis, use of valid methods for identified conditions, valid measurement for all participants, use of appropriate statistical analysis, and adequate response rate (≥60%). Each parameter was scored as 0 (yes) or 1 (no). Risk of bias was classified as low (0-2), moderate (3-4), or high (5-9).

### Outcome of Measurement

The primary outcomes of this study were the 12-month and lifetime prevalence of occupational exposure to BBFs. These prevalence rates were calculated by dividing the number of HCWs who experienced an occupational exposure by the total number of HCWs enrolled in the study and who were at risk of experiencing an occupational exposure to BBFs. Similarly, the reporting rate was calculated by dividing the number of cases reported or managed by the hospital focal point by the total number of cases reported by study participants

### Operational Definition

Occupational exposure in this report includes any splash exposure of BBFs into the eyes, nose, or mouth or non-intact skin, as well as exposure to BBFs through percutaneous injuries.

### Statistical Analysis

The researchers estimated the pooled prevalence of BBFs exposure using common- and random-effects meta-analysis models based on the Der Simonian and Laird approach. The existence of heterogeneity among the studies was assessed using the *I^2^* test statistic. Three categories were used to classified the degree of heterogeneity (*I^2^* index). These categories were either low (<25%), moderate (25-75%), or high (>75%) heterogeneity. The subgroup analysis was performed for the study year, the region, the sample size and the hospital level. The random-effects model was preferred if a significant heterogeneity between studies, was observed for the 12-month and lifetime BBFs exposure prevalence estimates.

A *p*-value<0.05 was used to declare significant heterogeneity. Finally, meta-regression analysis was used to assess the association between the prevalence of BBFs exposure and the study year, the region and the sample size. Data were analyzed using R Statistics version 4.3.3. Publication bias was assessed using funnel plots. Symmetrical large inverted funnels reflected the absence of publication bias. A narrative review of the significant risk factors was also performed.

## Results

A comprehensive literature review yielded a total of 539 records. After removal of 144 duplicate studies, 395 records remained. After reviewing their titles and abstracts, 319 reports were excluded for lack of relevance. The full texts of the remaining 16 records were then assessed for eligibility. Finally, 15 study reports met the eligibility criteria and were included in this systematic review and meta-analysis (Figure 1).

### Description of Studies

**Fig. 1.**
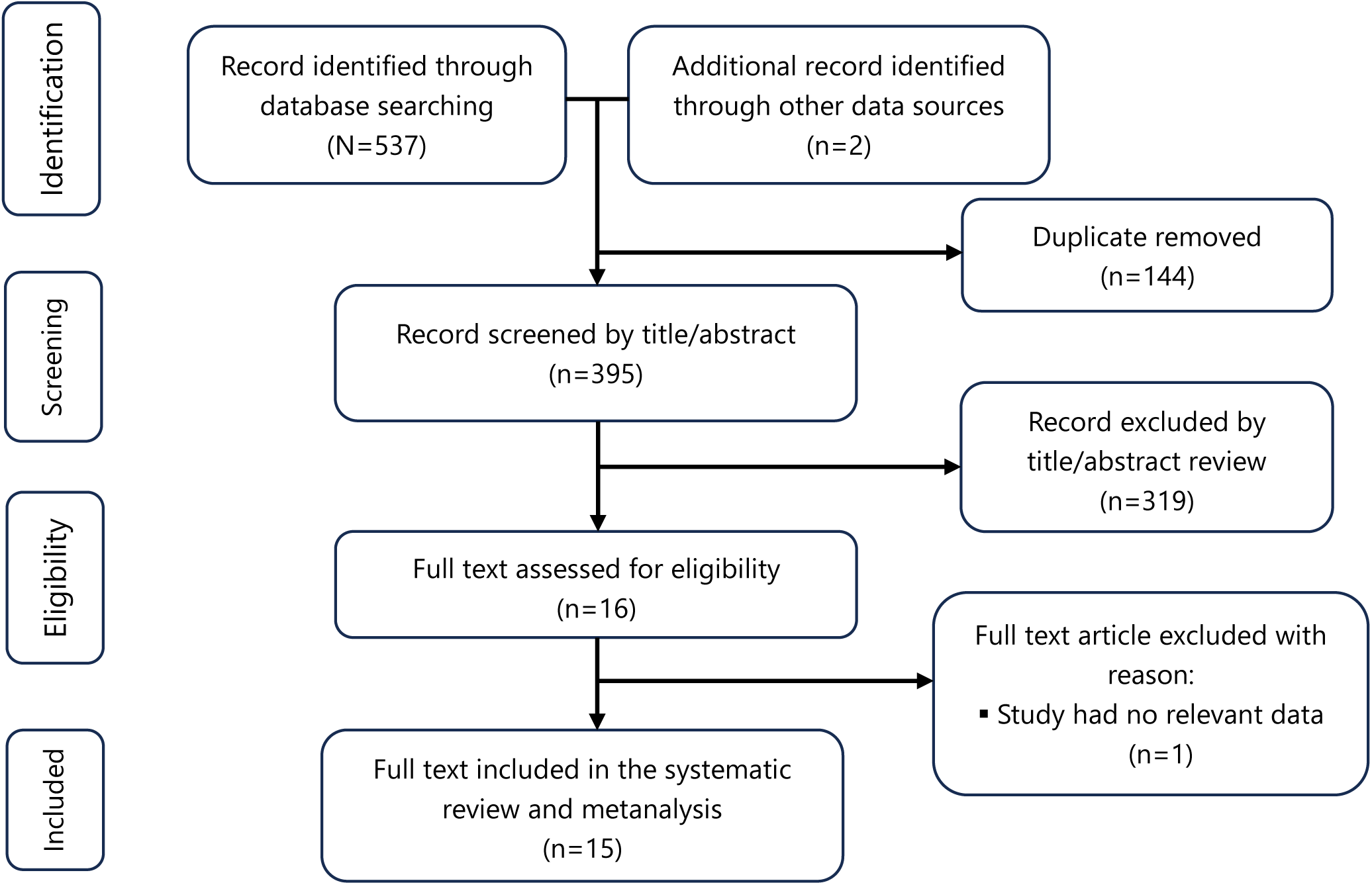
Flow diagram of the systematic review of studies related to occupational exposure among healthcare workers in Cameroon, 2010–2023

A total of 14 original articles and one unpublished research for total sample of 2613 participants were included to estimate the pooled prevalence, determine the reporting rate and review risk factors. The studies were conducted between 2012 and 2023. Most of the studies were conducted in the Centre Region and in central-level healthcare settings (Table 1).

**Table 1.** Characteristic of studies included to assess the pooled prevalence of occupational exposure among healthcare workers in Cameroon, 2010-2023.

| Author | Region | Year of study | Design | Setting | Study population | Sample size | Response rate (%) | Quality score |
| --- | --- | --- | --- | --- | --- | --- | --- | --- |
| Noubiap *et al.* [19] | Centre | 2012 | CS | Central level hospital<br>University | Medical student | 111 | 100 | 6 |
| Aminde *et al.* [13] | North-west | 2013 | CS | District hospital | HCWs | 80 | 94 | 7 |
| Domkam *et al.* [20] | Centre<br>East<br>Littoral<br>North-west<br>South-west | 2015 | CS | Intermediate and operational level hospitals | HCWs | 446 | 100 | 8 |
| Esum *et al.* [21] | Centre<br>South-west | 2019 | CS | District hospital | HCWs | 312 | 100 | 7 |
| Ngekeng *et al.* [22] | South-west | 2018 | CS | Intermediate and operational level hospital | HCWs | 281 | 100 | 7 |
| Nouetchougnou *et al.* [11] | Centre | 2013 | CS | Central level hospital | HCWs | 150 | 64.4 | 8 |
| Maka *et al.* [23] | Centre | 2016 | CS | District hospital | HCWs | 43 | 43 | 5 |
| Mbanya *et al.* [24] | Centre | 2010 | CS | Central level hospital | HCWs | 409 | 60.1 | 7 |
| Cheuyem *et al.* [3,10,25] | Centre | 2022 | CS | District hospital | HCWs | 217 | 78 | 8 |
| Takougang *et al.* [26] | Centre | 2021 | CS | Central level hospital | HCWs | 40 | 75.5 | 5 |
| Takougang *et al.* [4] | East | 2021 | CS | Intermediate level hospital | HCWs | 120 | 60 | 8 |
| Takougang *et al.* [12] | South-west | 2023 | CS | Intermediate level hospital | HCWs | 200 | 80 | 7 |
| Fotzo *et al.* [27] | Centre | 2022 | CS | Central level hospital | HCWs | 204 | 77 | 6 |

CS: Cross sectional study; HCW: Healthcare worker

### Prevalence of Occupational Exposure to BBFs

The random-effects model showed that the estimated overall pooled prevalence of 12-month exposure to BBFs among HCWs in Cameroon was 55.44% (95% CI: 41.20-69.68) with a highly significant level of heterogeneity (*I^2^*=97.5%; *p*<0.001). The pooled lifetime prevalence of exposure to BBFs using the random-effects model was 57.27% (95% CI: 42.43-72.10) with a significantly high level of heterogeneity (*I^2^*=97.7%; *p*<0.001) (Figures 2 and 3).

**Fig. 2.**
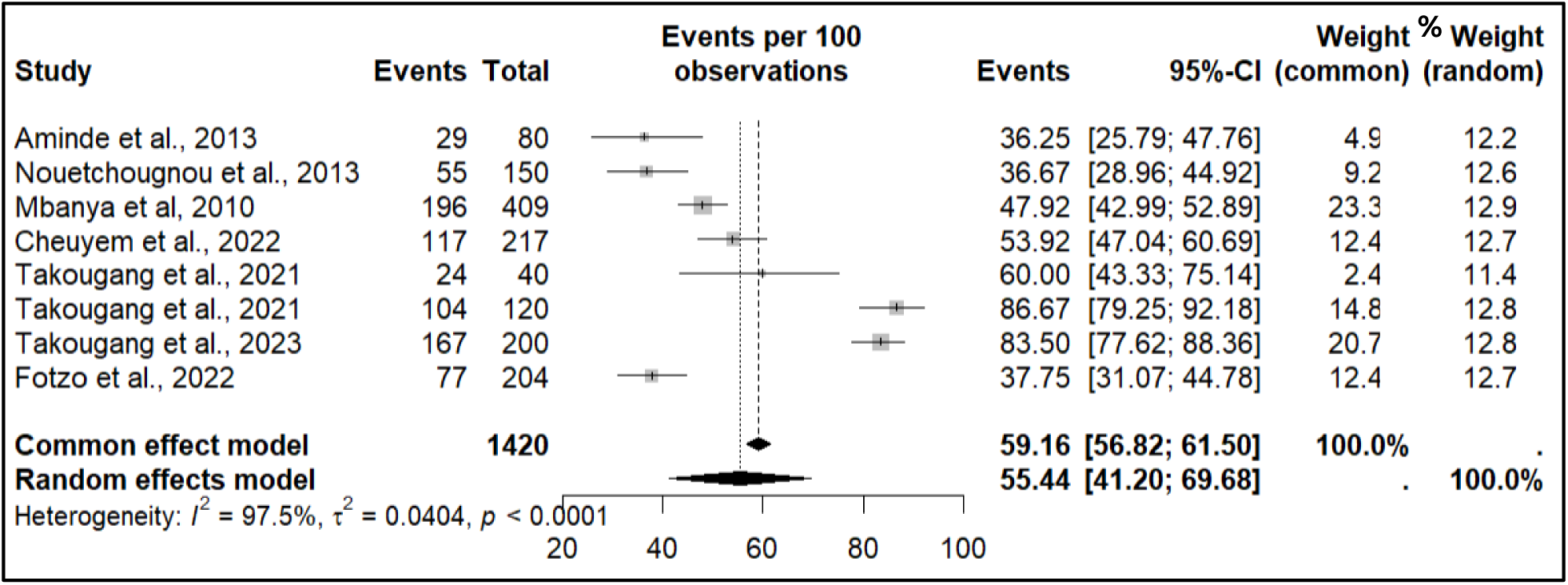
Annual estimates of the occupational exposure to body fluids among healthcare workers in Cameroon, 2010-2023

**Fig. 3.**
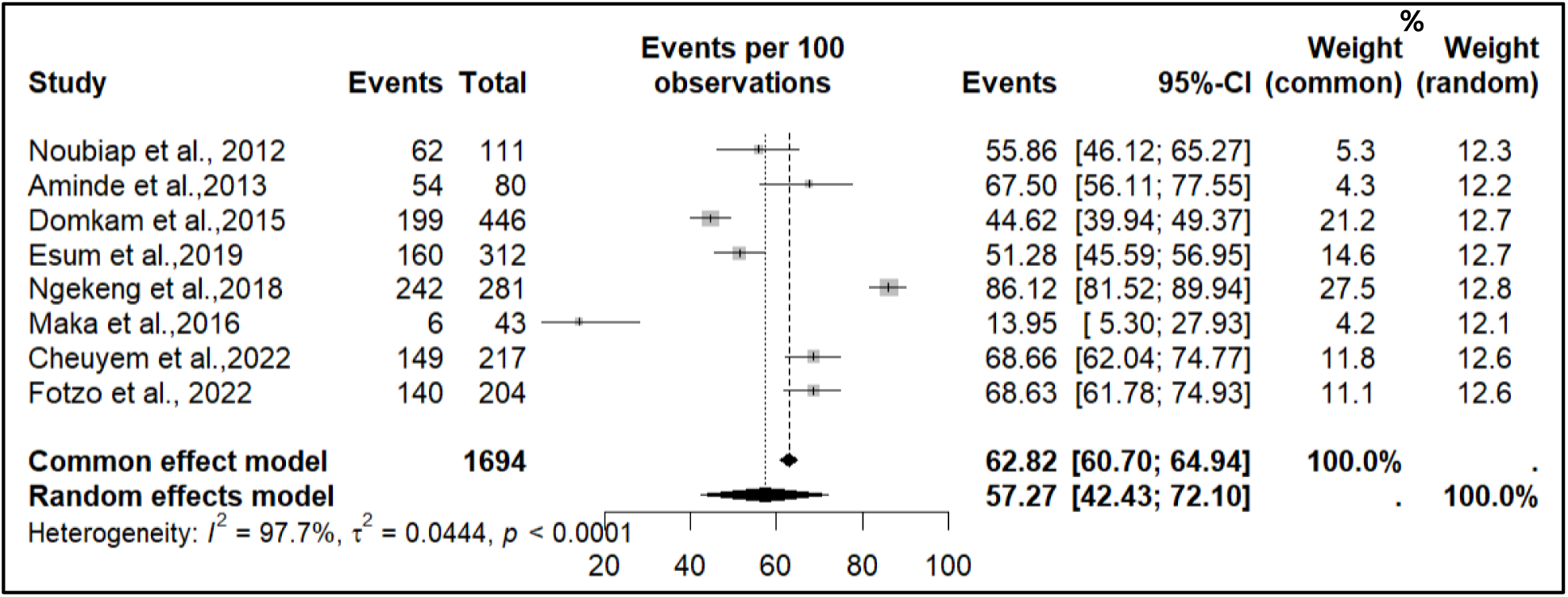
Life course estimates of the occupational exposure to body fluids among healthcare workers in Cameroon, 2010-2023

### Heterogeneity and Subgroup Analysis

The random-effects model was preferred because of the highly significant heterogeneity between studies for both 12-month and lifetime BBFs exposure prevalence estimates (Figure 2 and 3).

The highest 12-month pooled prevalence was observed in intermediate level health facilities (84.73%; 95% CI: 85.55-88.50), in Regions other than the Centre (70.87%; 95% CI: 37.26-95.13) and for studies conducted from 2017 to 2023 (65.63%; 95% CI: 45.73-83.06) and the lowest was reported from studies conducted from 2010 to 2016 (41.13%; 95% CI: 33.11-49.40). This meta-analysis also found that the lifetime prevalence of BBFs exposure was the highest for other Region namely the North-west and South-west Regions (77.96%; 95% CI: 57.39-93.19) and the lowest observed in studies with sample size less than 200 45.9% 95% CI: 14.81-77.50) or studies conducted from 2010 to 2016 (45.11%; 95% CI: 23.10-68.16) (Table 2).

**Table 2.**
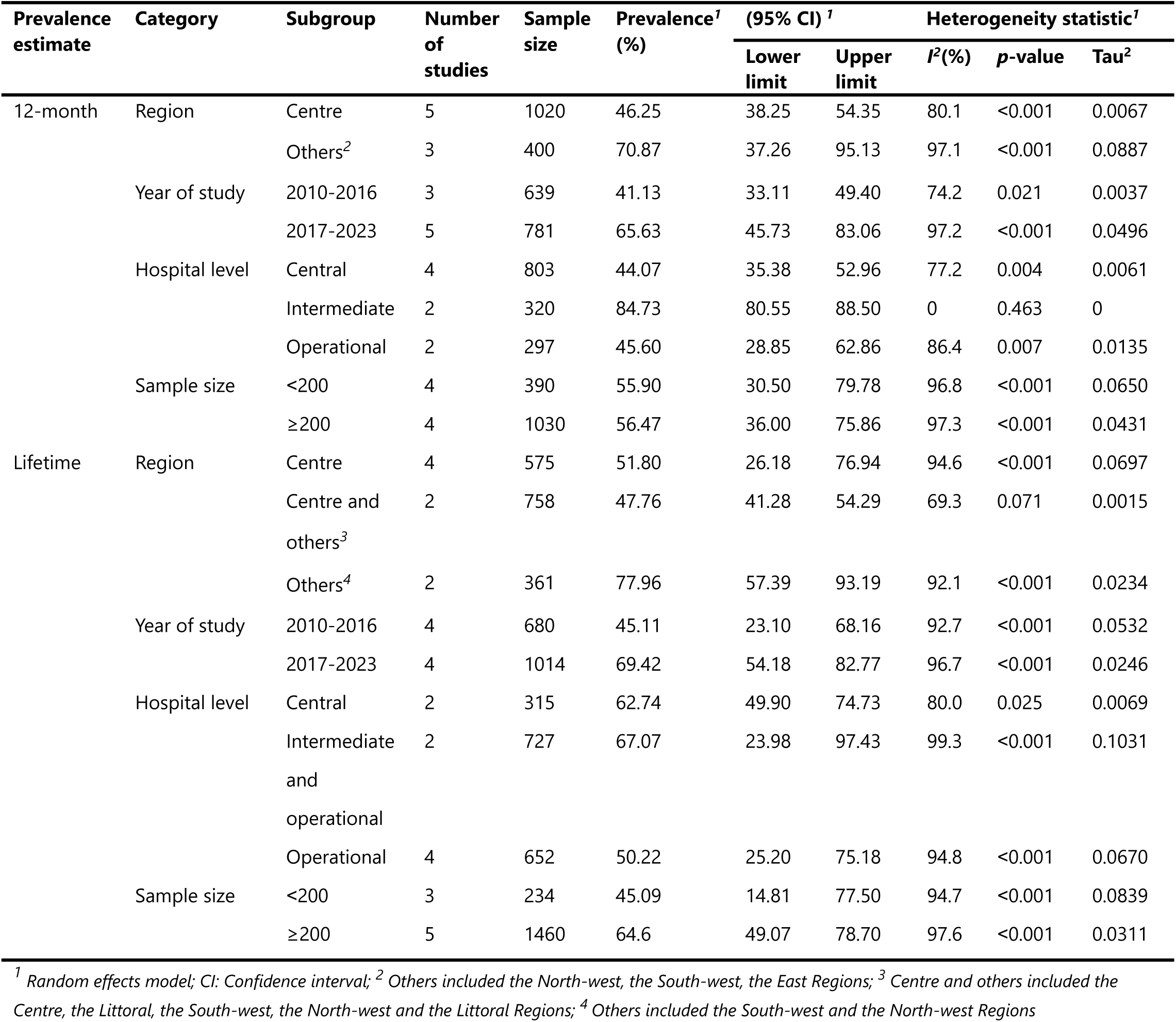
Subgroup metanalysis of prevalences estimates of occupation exposure among healthcare workers in Cameroon, 2010-2023.

| Prevalence estimate | Category | Subgroup | Number of studies | Sample size | Prevalence<sup>1</sup> (%) | (95% CI) <sup>1</sup> |  | Heterogeneity statistic<sup>1</sup> |  |  |
| --- | --- | --- | --- | --- | --- | --- | --- | --- | --- | --- |
|  |  |  |  |  |  | Lower limit | Upper limit | I<sup>2</sup>(%) | p-value | Tau<sup>2</sup> |
| 12-month | Region | Centre | 5 | 1020 | 46.25 | 38.25 | 54.35 | 80.1 | <0.001 | 0.0067 |
|  |  | Others<sup>2</sup> | 3 | 400 | 70.87 | 37.26 | 95.13 | 97.1 | <0.001 | 0.0887 |
|  | Year of study | 2010-2016 | 3 | 639 | 41.13 | 33.11 | 49.40 | 74.2 | 0.021 | 0.0037 |
|  |  | 2017-2023 | 5 | 781 | 65.63 | 45.73 | 83.06 | 97.2 | <0.001 | 0.0496 |
|  | Hospital level | Central | 4 | 803 | 44.07 | 35.38 | 52.96 | 77.2 | 0.004 | 0.0061 |
|  |  | Intermediate | 2 | 320 | 84.73 | 80.55 | 88.50 | 0 | 0.463 | 0 |
|  |  | Operational | 2 | 297 | 45.60 | 28.85 | 62.86 | 86.4 | 0.007 | 0.0135 |
|  | Sample size | <200 | 4 | 390 | 55.90 | 30.50 | 79.78 | 96.8 | <0.001 | 0.0650 |
|  |  | ≥200 | 4 | 1030 | 56.47 | 36.00 | 75.86 | 97.3 | <0.001 | 0.0431 |
| Lifetime | Region | Centre | 4 | 575 | 51.80 | 26.18 | 76.94 | 94.6 | <0.001 | 0.0697 |
|  |  | Centre and others<sup>3</sup> | 2 | 758 | 47.76 | 41.28 | 54.29 | 69.3 | 0.071 | 0.0015 |
|  |  | Others<sup>4</sup> | 2 | 361 | 77.96 | 57.39 | 93.19 | 92.1 | <0.001 | 0.0234 |
|  | Year of study | 2010-2016 | 4 | 680 | 45.11 | 23.10 | 68.16 | 92.7 | <0.001 | 0.0532 |
|  |  | 2017-2023 | 4 | 1014 | 69.42 | 54.18 | 82.77 | 96.7 | <0.001 | 0.0246 |
|  | Hospital level | Central | 2 | 315 | 62.74 | 49.90 | 74.73 | 80.0 | 0.025 | 0.0069 |
|  |  | Intermediate and operational | 2 | 727 | 67.07 | 23.98 | 97.43 | 99.3 | <0.001 | 0.1031 |
|  |  | Operational | 4 | 652 | 50.22 | 25.20 | 75.18 | 94.8 | <0.001 | 0.0670 |
|  | Sample size | <200 | 3 | 234 | 45.09 | 14.81 | 77.50 | 94.7 | <0.001 | 0.0839 |
|  |  | ≥200 | 5 | 1460 | 64.6 | 49.07 | 78.70 | 97.6 | <0.001 | 0.0311 |

<sup>1</sup> Random effects model; CI: Confidence interval; <sup>2</sup> Others included the North-west, the South-west, the East Regions; <sup>3</sup> Centre and others included the Centre, the Littoral, the South-west, the North-west and the Littoral Regions; <sup>4</sup> Others included the South-west and the North-west Regions

### Reporting Pattern of Occupational Exposure to BBFs

The random-effects model showed that the estimated overall pooled reporting rate in Cameroon was 54.72% (95% CI: 43.50-65.95) with a highly significant level of heterogeneity (*I^2^*=84.9%; *p*<0.001) (Figure 4).

**Fig. 4.**
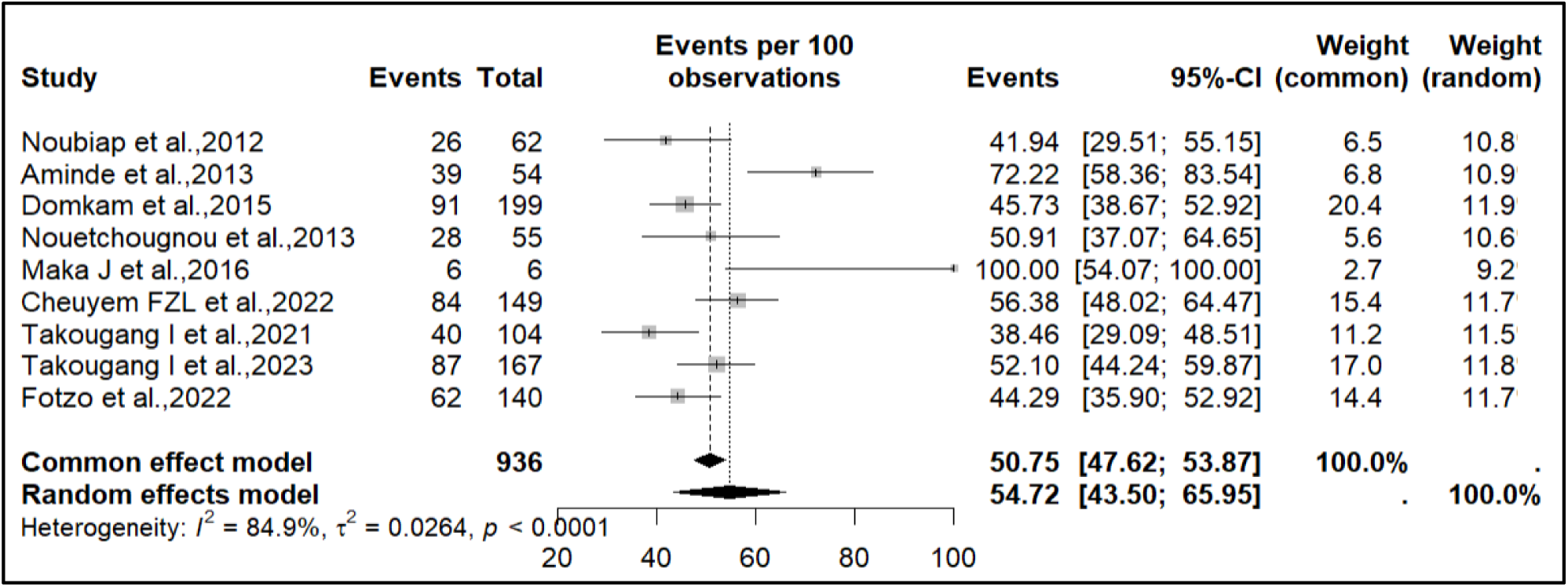
Reporting estimates of occupational exposures cases among healthcare workers in Cameroon, 2010-2023

Visual inspection of the funnel plot revealed a relatively symmetrical distribution of studies assessing the lifetime prevalence of occupational exposure to BBFs and the reporting profile. This symmetry suggests that the studies are scattered around the pooled estimate in a fairly even manner, with no obvious evidence of publication bias or small-study effects. However, the asymmetry was observed among studies assessing the 12-month prevalence reflecting potential publication bias (Figure 5).

**Fig. 5.**
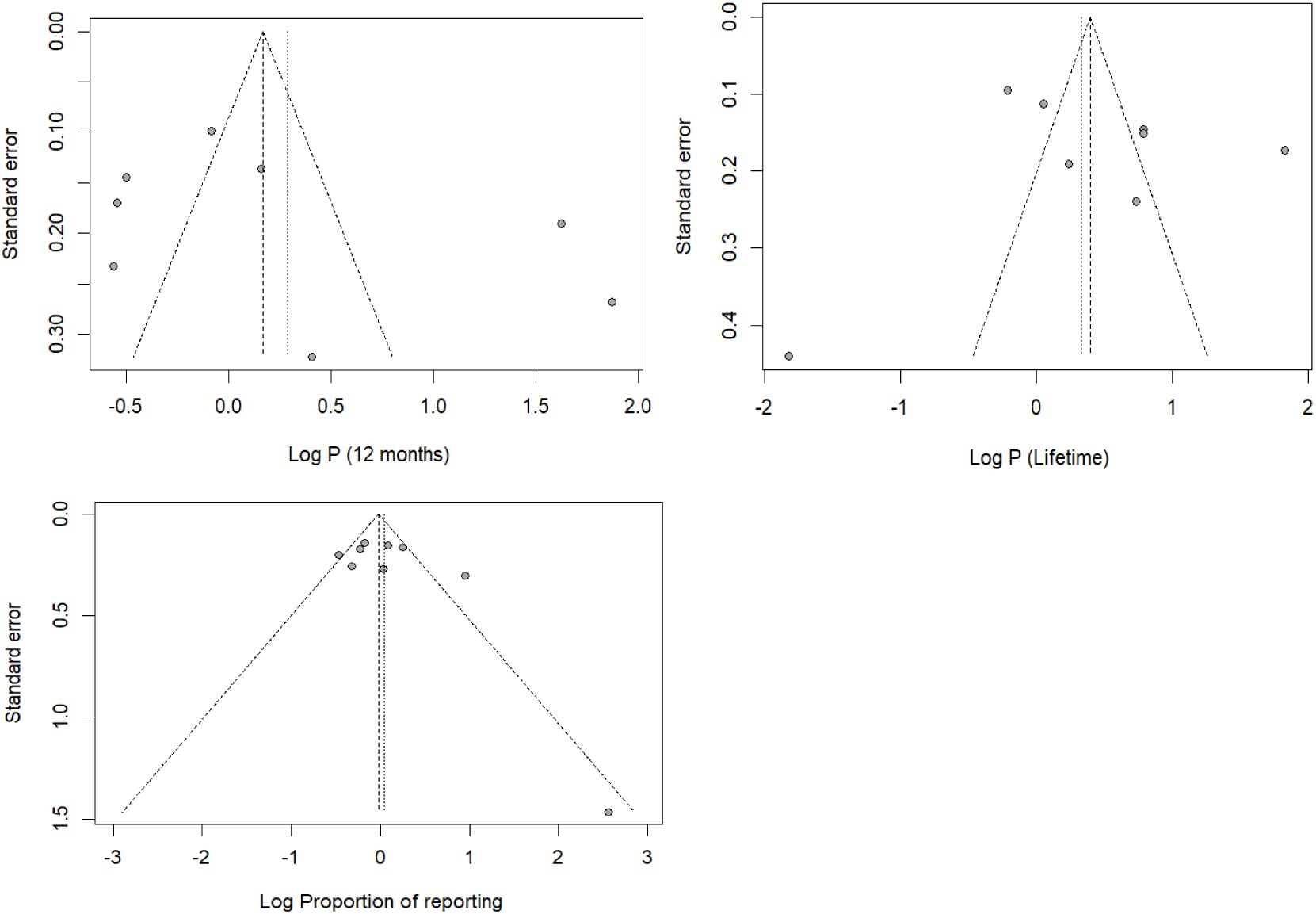
Funnel plot with pseudo 95% confidence limits of studies included

### Meta-Regression Analysis

The meta-regression analysis showed that the observed heterogeneity in the 12-month and lifetime prevalence of occupational exposure to BBFs was not due to differences in study year, sample size or region (Table 3).

**Table 3.**
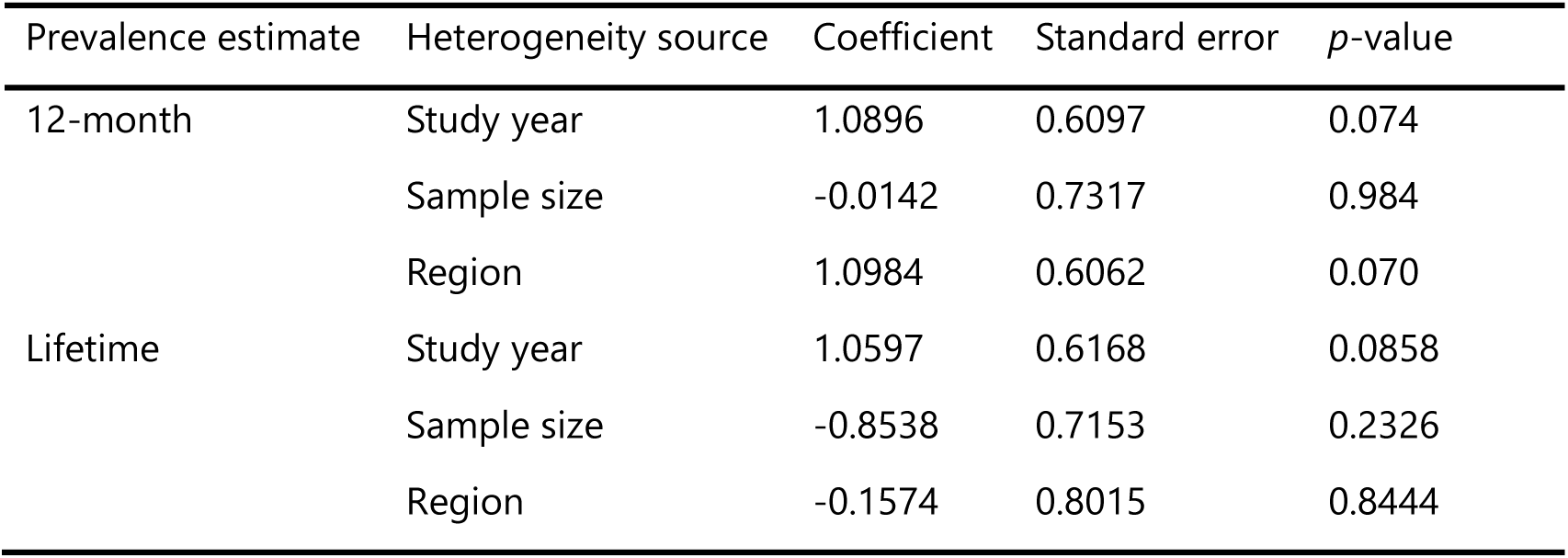
Meta-regression of factor influencing the heterogeneity of the prevalence of occupational exposure among healthcare worker in Cameroon, 2010-2023.

**Table 4.** Parameters associated with occupational exposure to blood and other body fluids among healthcare workers in Cameroon, 2010-2023.

| Author | Study design | Study population | Setting | Region | Results |
| --- | --- | --- | --- | --- | --- |
| Cheuyem *et al.* [25] | CS | HCWs | Hospital | Centre | HCWs aged less than 35 years (AOR 2.37; 95% CI: 1.09-5.22), with less than 12 years of education (AOR 0.34; 95% CI: 0.11-0.91), working in a surgical unit (AOR 3.11; 95% CI: 1.33-7.38), and recycling scalp blades for reuse (AOR 7.56; 95% CI: 2.44-25.9) were at significantly higher risk of percutaneous injury. HCWs working in the surgical unit (AOR 4.72; 95% CI: 1.95-12.3), in the obstetric unit (AOR 3.16; 95% CI: 1.30-8.13), those who did not receive refresher training (AOR 2.88; 95% CI: 1.48-5.82), and those who were fully vaccinated against HVB (AOR 6.47; 95% CI: 2.25-20.5) were at significantly higher risk of splash exposure. |
| Esum *et al.* [21] | CS | HCWs | Hospital | Centre and South-west | HCWs with tertiary education (AOR 0.34; 95% CI: 0.16-0.75), or working in a surgical unit (AOR 3.11; 95% CI: 1.33-7.38) were significantly more at risk of percutaneous exposure to BBFs. |
| Takougang *et al.* [4] | CS | HCWs | Hospital | East | Midwives (AOR 65.9; 95% CI: 8.49-1428) and cleaners (AOR 14.7; 95% CI: 1.44-386) had a significantly higher risk of occupational exposure to BBFs. |
| Takougang *et al.* [12] | CS | HCWs | Hospital | South-west | HCWs from the medical (AOR 5.95; 95% CI: 1.77-24.3), surgical (AOR 3.74; 95% CI: 1.06-15.7), and pediatric (AOR 10.9; 95% CI: 2.07-74.3) wards were significantly more at risk for percutaneous injury compared to those from the laboratory unit. Female HCWs (AOR 2.86; 95% CI: 1.23-7.18), those working in the laboratory (AOR 13.3; 95% CI: 2.51-109), obstetrics (AOR 22.6; 95% CI: 4.3-186), dental service (AOR 26.3; 95% CI: 2.91-361) were significantly more at risk of splashes compared to those working in the emergency department. HCWs who had not received refresher training on ICP (AOR 2.41; 95% CI: 5.01-0.015) were significantly more likely to underreport occupational exposure to BBFS. |

CS: Cross-sectional study; HCW: Healthcare worker; AOR: Adjusted Odds Ratio; CI: Confidence interval; BBF: Blood and other body fluid, ICP: Infection control and prevention

### Narrative Review of Factors Associated with Occupational Exposures to BBFs

Various factors were identified as associated with the occurrence of occupational exposure to BBFs. Such factors included the age, the educational level, the working unit (surgical, obstetrical wards), professional status (midwifes, cleaners), lack of refresher training in infection control and prevention measures.

## Discussion

Health information is needed to make evidence-based decisions about the health and well-being of HCWs, who are one of the main pillars of the health system [14]. This first systematic review and meta-analysis aimed to estimate the lifetime and 12-month prevalence of occupational exposure to BBFs among healthcare workers (HCWs) in Cameroon, providing evidence of the burden of this phenomenon among this professional group. We included a total of 15 study reports that investigated the prevalence of BBFs exposure and demonstrated a high burden of occupational exposure to BBFs among HCWs in Cameroon.

More than half of HCWs experience an occupational exposure either in a lifetime (57.27%) or 12-month (55.44%) period. Occupational exposure to BBFs is a major public health problem in Cameroon, as in many other countries of the world. The results of this study show that HCWs in Cameroon are at high risk of infection from BBFs, particularly HIV, hepatitis B, and hepatitis C. These findings are consistent with studies conducted in other sub-Saharan African and Asian countries, where rates of occupational exposure to BBFs are also high [7,28–30]. Reports from worldwide systematic review and metanalysis depict similar trends [31].

The study results suggest that HCWs in Cameroon are at high risk of exposure to BBFs, especially in intermediate level health facilities and in Regions other than the Center. Health facilities in the Centre Region, particularly the referral hospitals where most of the included studies were conducted, may have benefited from better training, provision of personal protective equipment, and a safer working environment compared to health facilities in other Regions. In the fight against this medical hazard, this evidence can be used to direct efforts and resources to where the burden of the phenomenon is higher. Especially in a resource-limited country.

This study showed an increase in both 12-month and lifetime prevalence of occupational exposure from 2010 to 2023. This may be due to the deficiencies in the implementation of adequate occupational safety and health policies and guidelines to address this health problem in the country. Such policies should ensure the implementation of medical education and training on infection control practices and occupational safety in healthcare settings, the availability of personal protective equipment and the adequate waste management practices [8]. In addition, the increased patient load of HCWs in a context of a limited health workforce might have influenced this trend [17]. In this regard, studies from Cameroon and Ethiopia report that fatigue and stress due to high workload were commonly cited factors favoring the occurrence of occupational exposure to BBFs [25,32,33].

## Limitations

The asymmetry observed in the funnel plot suggests that smaller studies with more extreme results may be overrepresented in the literature, potentially leading to an overestimation of the true prevalence of occupational exposure to BBFs. This limitation highlights the need for future studies to use larger sample sizes and more rigorous reporting methods. In addition, the included studies may be subject to recall bias, which could affect the accuracy of the results. Furthermore, the geographical representation of the studies was limited, with no studies from certain regions of Cameroon, such as the northern part of the country. Despite these limitations, this systematic review and meta-analysis used robust methods to critically appraise and synthesize the available evidence. The use of internationally accepted tools and the inclusion of both published and unpublished studies helped to minimize publication bias and address the underreporting of negative results.

## Conclusions

HCWs in Cameroon face a significant risk of occupational exposure to BBFs, with more than half experiencing exposure in their lifetime or within a 12-month period. This is a major public health concern, particularly for HIV, hepatitis B and hepatitis C infections transmission. The findings highlight the need for targeted interventions, including improved infection control practices, adherence to standard precautions, and enhanced occupational safety and health policies.

## Data Availability

All data produced in the present work are contained in the manuscript

## Abbreviations

BBFs: Blood and other Body Fluids
CI: Confidence Interval
CS: Cross Sectional Study
HCW: Healthcare Worker
HIV: Human Immunodeficiency Virus
ICP: Infection Control and Prevention
MeSH: Medical Subject Headings
PRISMA: Preferred Reporting Items for Systematic Reviews and Meta-Analysis

## Declarations

### Author contributions

FZLC conceived the original idea of the study. FZLC and DCMH conducted the literature search. FZLC, MCE, and CM selected the studies, extracted the relevant information, and synthesized the data. FZLC performed the analyses and wrote the first draft of the manuscript. All authors critically reviewed and revised successive drafts of the manuscript. All authors read and approved the final manuscript.

### Ethical Approval Statement

Not applicable

### Consent for publication

Not applicable.

### Availability of data and materials

All data generated or analyzed during this study are included in this published article.

### Competing interests

All authors declare no conflicts of interest and have approved the final version of the article.

### Funding source

This research did not receive any specific grant from funding agencies in the public, commercial or not-for-profit sectors.

## Notes

### Competing Interest Statement

The authors have declared no competing interest.

### Funding Statement

This study did not receive any funding

### Author Declarations

Source data were openly available before the initiation of the study. Links to included research article are available in the reference of the paper. https://doi.org/10.1186/s12913-024-10855-x https://www.opastpublishers.com/peer-review/accidental-exposure-to-body-fluids-among-healthcare-workers-in-a-referral-hospital-in-the-securitychallenged-region-of-s-7302.html http://dx.doi.org/10.34297/AJBSR.2023.19.002628 https://www.primescholars.com/articles/infection-risk-perception-reporting-and-postexposure-management-of-occupational-injuries-among-healthcare-workers-in-district-hosp-123143.html https://bmcmededuc.biomedcentral.com/articles/10.1186/1472-6920-13-148 https://journalmrji.com/index.php/MRJI/article/view/56 https://journalmrji.com/index.php/MRJI/article/view/56 https://doi.org/10.21522/TIJPH.2013.05.04.Art029 https://doi.org/10.11604/pamj.2018.29.158.14073 http://www.biomedcentral.com/1756-0500/9/94 https://www.mdpi.com/1660-4601/7/5/2085

### Summary of Updates

The list of authors of the system has been revised to align with the one available in the paper.

